# Coronary revascularisation in people with multiple long-term conditions: a systematic review and meta-analysis

**DOI:** 10.64898/2026.09.04.26362027

**Authors:** Ann Cheng, Ali Alhashimi, Chai Jin Lim, Sxe Chang Cheong, Hunain Shahbaz, Riccardo Abbasciano, Maria Pufulete, Gavin J Murphy

## Abstract

**Introduction:** People presenting for multivessel coronary revascularisation commonly have multiple long-term conditions that influence prognosis. The aim of this study was to review the evidence for revascularisation choices in these groups.

**Methods:** Randomised trials of Coronary Artery Bypass Grafting (CABG) versus Percutaneous Coronary Intervention (PCI) reporting outcomes by pre-specified comorbidities were included. Treatment effects were estimated using inverse variance random effects models and presented as Risk Ratios (95% Confidence Intervals). The certainty of evidence for outcomes was assessed using Grading of Recommendations Assessment, Development and Evaluation (GRADE).

**Results:** 19 studies from nine RCTs with 10,594 participants were included. Reported baseline comorbidities included diabetes (49%), CKD (25%), LVSD (9%), female sex (14%), and older age (40%). Trials were unblinded, resulting in some bias concerns. For people with diabetes and CKD, CABG reduced the risks of all-cause death and repeat revascularisation, with no treatment effect on MI or stroke, versus PCI. There was no difference in mortality or stroke for female sex, or older age. No data was reported for people with peripheral vascular disease or frailty. Patient reported outcomes favoured CABG in people with diabetes. GRADE showed that the certainty of the evidence was generally Low to Very Low with the exception of people with diabetes (Moderate Certainty).

**Conclusion:** There is a knowledge gap with respect to the risks and benefits of CABG versus PCI in people with common comorbidities.

**Registration:** PROSPERO CRD420251032665

## INTRODUCTION

Coronary artery disease (CAD) is the leading cause of death worldwide, accounting for 9.1 million deaths in 2021.[1] Treatment guidelines recommend myocardial revascularisation in symptomatic multivessel coronary artery disease to reduce death and major adverse cardiovascular events. Based on RCT evidence, guidelines recommend revascularisation with either CABG or PCI based on the severity and extent of coronary artery disease, and the presence or absence of diabetes and ischaemic left ventricular dysfunction (iLVSD).[2–4]

Coronary artery disease commonly presents as one of multiple long-term conditions (MLTC), defined as the co-existence of two or more chronic (> 1 year duration) conditions.[5] Other long-term conditions such as chronic kidney disease, peripheral vascular disease, and heart failure, or frailty, are important considerations in treatment decisions for coronary artery disease, as they strongly influence clinical outcomes.[6] However, people with these conditions were typically excluded or under-represented in previous trials of CABG versus PCI, and guidelines make no references to these groups beyond recommendations for PCI in people at high surgical risk.[7] This knowledge gap is associated with variation in treatment choices; the proportion of people with MLTC undergoing multivessel coronary revascularisation with CABG ranges from 25% to 82% across regions in England.[7] This is also a major contributor to health inequality, as MLTC are over-represented in underserved populations including female sex, black ethnicity, and living in areas of social deprivation.[8]

The aim of this systematic review was to examine the effectiveness of CABG versus PCI in people with MLTC. To rationalise the scope of this work, MLTC populations of interest included diabetes, chronic kidney disease (CKD), left ventricular systolic dysfunction (LVSD), peripheral vascular disease (PVD), and frailty; female and people aged > 75 years are often considered as MLTC and are also included.

## METHODS

### Protocol and registration

The study protocol was prospectively registered; PROSPERO CRD420251032665. The results are reported as per the Preferred Reporting Items for Systematic Reviews and Meta-Analyses (PRISMA) guideline.[9] Informed consent was not required as this was a study-level systematic review and no human subjects were involved.

### Information sources & search strategy

One reviewer (AC) performed the search using Ovid Medline, EMBASE, CINAHL, CENTRAL (Cochrane Central Register of Controlled Trials), and PsycINFO on 10 January 2025. Limits applied, where possible, were publication year after 2000 and English language only. The reference sections of previously published systematic reviews were also screened for additional eligible articles. Key search terms used included Medical Subject Headings (MeSH) and associated keywords for coronary revascularisation including variations of PCI and CABG, coronary artery disease, and multiple long-term conditions; an example search strategy is available in Supplementary Materials.

### Eligibility criteria

RCTs comparing PCI and CABG reporting any MLTC of interest; diabetes, CKD, LVSD, PVD, frailty, female, and age > 75 years were included. Observational studies, including registry data, single-centre outcomes, or those without comparator groups, were deemed at high risk of bias and excluded. Studies recruiting prior to 2000 were excluded pragmatically due to significant advances in PCI after the introduction of drug eluting stents.

### Study selection

Search records were collated, and duplicates were removed. Three reviewers (AA, CJL, SCC) independently screened all titles and abstracts sequentially for eligibility using Rayyan [10]; a pilot of 25 records were screened by all reviewers to ensure concordance. An additional reviewer (AC) checked at least 10% random sample for accuracy due to the large number of study records.[11] Full text of potentially eligible studies underwent final assessment according to the eligibility criteria in duplicates. Any discrepancies were resolved by discussion between the reviewers.

### Data extraction

Data was extracted in duplicate by 4 reviewers (AC, AA, CJL, HS) in Microsoft Excel[12] using a piloted extraction form; any discrepancies were resolved by discussion between the reviewers. Where data was available at multiple time points (e.g. revascularisation outcome for diabetes at 1, 3, and 5 years), the study reporting the longest follow-up was included.

### Data items

The primary outcome was all-cause mortality. Secondary outcomes included myocardial infarction (MI), stroke, repeat revascularisation, Major Adverse Cardiac and Cerebrovascular Events (MACCE), and quality of life, as defined by individual studies. Additional data items included the MLTC of interest and its definition used in respective studies, main trial population, whether MLTC outcome was pre-specified or post hoc, and participant demographics.

### Risk of bias assessment

Methodological quality of the included studies was assessed using the Revised Cochrane risk-of-bias tool for randomised trials (RoB2).[13, 14]

### Synthesis methods and statistical analysis

Dichotomous data were reported as risk ratios (RR) with 95% confidence intervals (CI), and continuous data as mean difference (MD) with 95% CIs. Heterogeneity was measured using I^2^ statistics in each analysis; high heterogeneity was defined as >80%. Subgroup analysis by MLTC was performed using an inverse variance random effects model in view of the clinical and methodological heterogeneity intrinsic to investigations of these interventions. Analyses and forest plot generation were carried out using RevMan version 5.4.1.[15] The certainty of evidence for outcomes was assessed using the Grading of Recommendations Assessment, Development and Evaluation (GRADE) framework.[16]

## RESULTS

### Eligible studies

Searches identified a total of 13,978 titles and abstracts, of which 19 full-text articles representing 9 RCTs comparing CABG versus PCI met the eligibility criteria and were included in the review (Figure 1). Six RCTs (ARTS1, ARTS2, AWESOME, BARI, MASS II, SoS) were excluded due to recruitment taking place prior to 2000; and 2 RCTs (Boudriot et al, LE MANS) were excluded due to not reporting any MLTC or subgroup analysis. Additionally, EXCEL reported outcomes for people with PVD and people aged > 75 years in two separate abstracts; however, no full text publications were subsequently identified.

**Figure 1.**
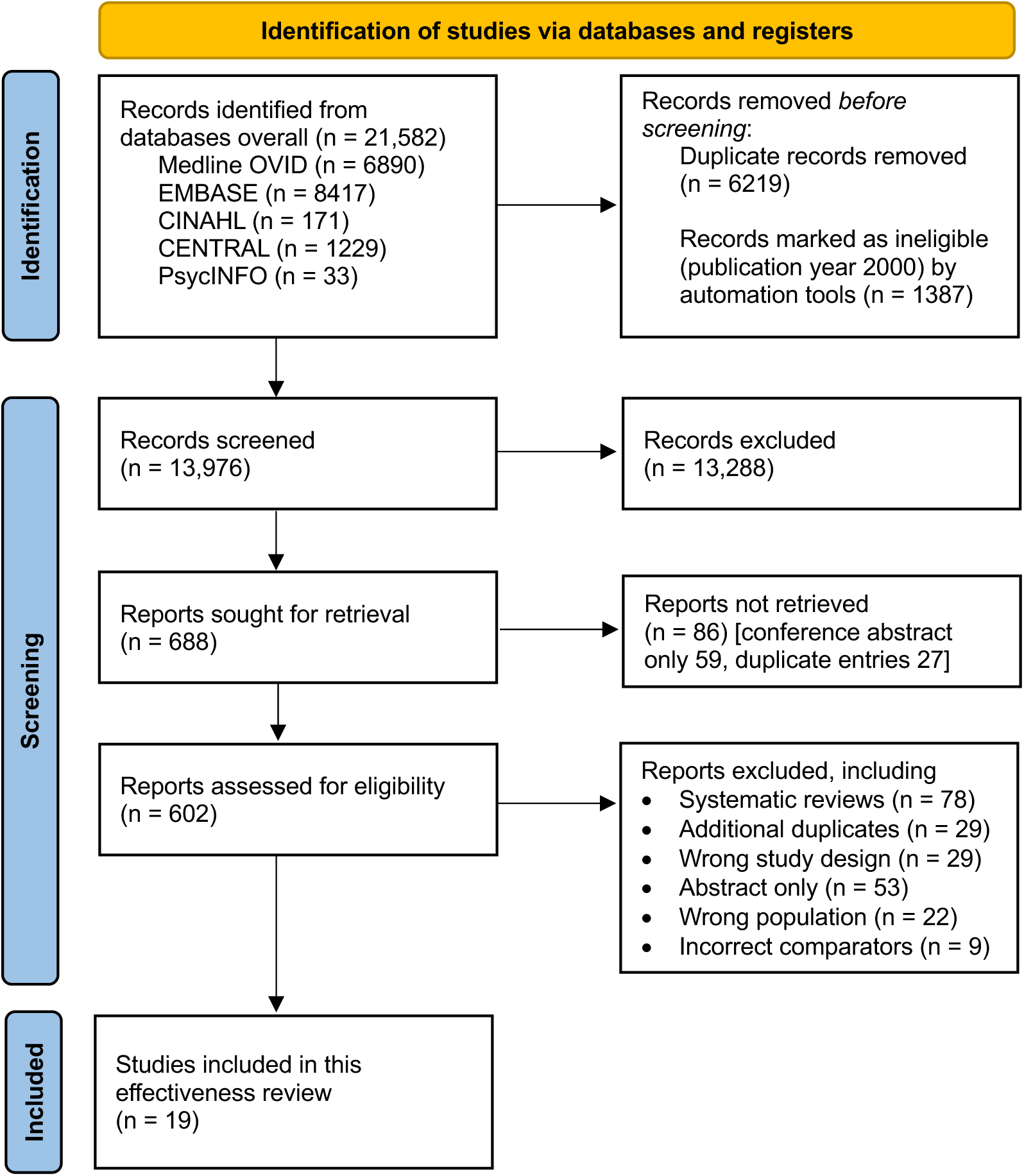
Summary of the search strategy process. Please note the diagram is representative of the wider ongoing mixed methods systematic review, except the included studies (n = 19)

### Study characteristics

A total of 5250 participants had PCI, and 5344 participants had CABG in 9 RCTs. The average age was 64 years and a quarter were female. 4 RCTs were international; 3 were regional; and 2 were limited to a single country. 3 RCTs included diabetes as an inclusion criterion; no RCT included any other LTC as an inclusion criterion. In contrast, exclusion criteria included congestive heart failure or LVEF < 30% in 5 trials, abnormal creatinine level or dialysis dependence in 2 trials, and an age cut-off above 80 years old in 1 trial. Pre-specified sub-studies or subgroup analyses addressing LTC for each RCT are described in Table 1.

**Table 1.**
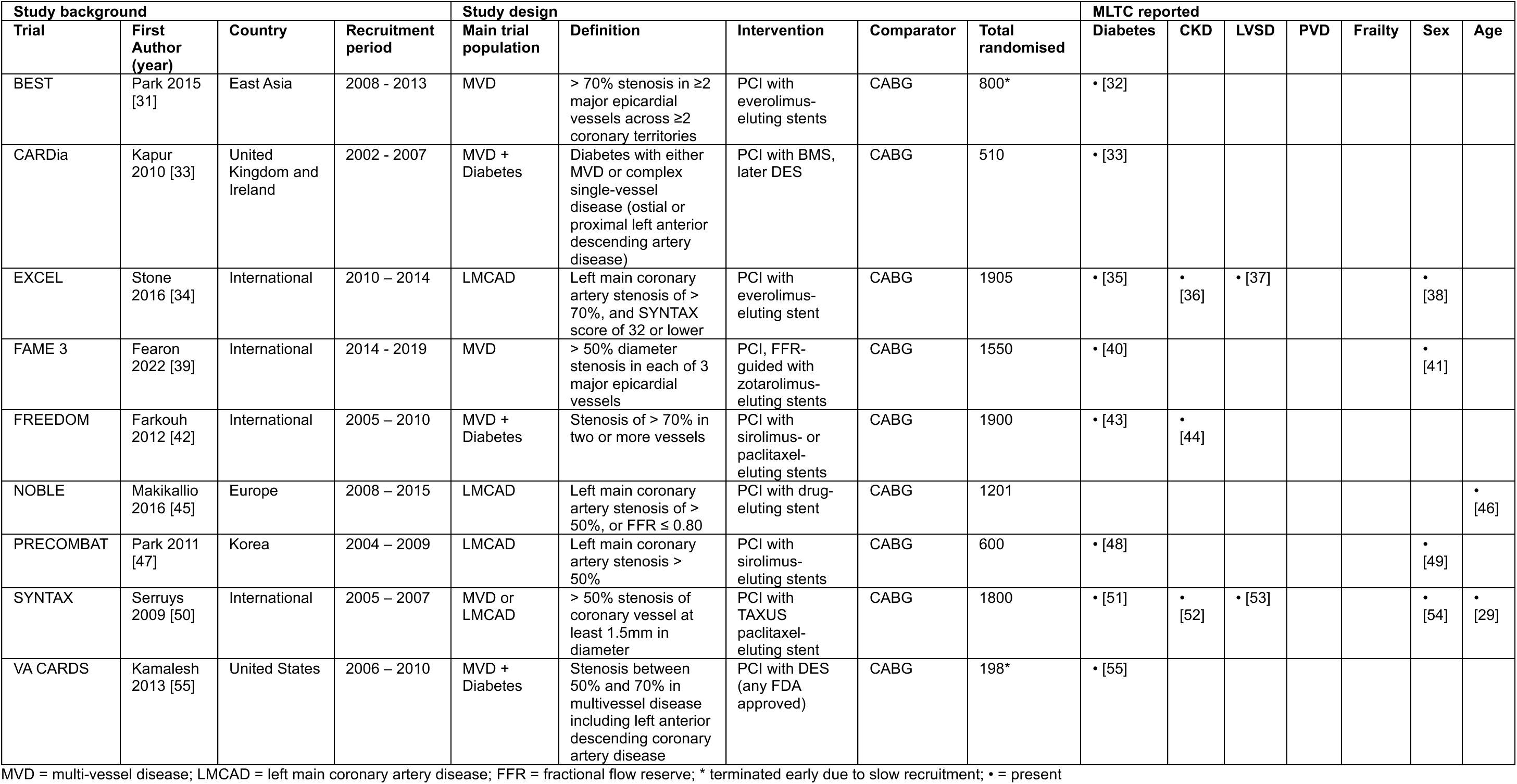
Study characteristics.

| Study background |  |  |  | Study design |  |  |  |  | MLTC reported |  |  |  |  |  |  |
| --- | --- | --- | --- | --- | --- | --- | --- | --- | --- | --- | --- | --- | --- | --- | --- |
| Trial | First Author (year) | Country | Recruitment period | Main trial population | Definition | Intervention | Comparator | Total randomised | Diabetes | CKD | LVSD | PVD | Frailty | Sex | Age |
| BEST | Park 2015 [31] | East Asia | 2008 - 2013 | MVD | > 70% stenosis in $\geq 2$ major epicardial vessels across $\geq 2$ coronary territories | PCI with everolimus-eluting stents | CABG | 800* | • [32] | | | | | | |
| CARDia | Kapur 2010 [33] | United Kingdom and Ireland | 2002 - 2007 | MVD + Diabetes | Diabetes with either MVD or complex single-vessel disease (ostial or proximal left anterior descending artery disease) | PCI with BMS, later DES | CABG | 510 | • [33] |  |  |  |  |  |  |
| EXCEL | Stone 2016 [34] | International | 2010 – 2014 | LMCAD | Left main coronary artery stenosis of > 70%, and SYNTAX score of 32 or lower | PCI with everolimus-eluting stent | CABG | 1905 | • [35] | • [36] | • [37] |  |  | • [38] |  |
| FAME 3 | Fearon 2022 [39] | International | 2014 - 2019 | MVD | > 50% diameter stenosis in each of 3 major epicardial vessels | PCI, FFR-guided with zotarolimus-eluting stents | CABG | 1550 | • [40] |  |  |  |  | • [41] |  |
| FREEDOM | Farkouh 2012 [42] | International | 2005 – 2010 | MVD + Diabetes | Stenosis of > 70% in two or more vessels | PCI with sirolimus- or paclitaxel-eluting stents | CABG | 1900 | • [43] | • [44] |  |  |  |  |  |
| NOBLE | Makikallio 2016 [45] | Europe | 2008 – 2015 | LMCAD | Left main coronary artery stenosis of > 50%, or FFR $\leq 0.80$ | PCI with drug-eluting stent | CABG | 1201 | | | | | | | • [46] |
| PRECOMBAT | Park 2011 [47] | Korea | 2004 – 2009 | LMCAD | Left main coronary artery stenosis > 50% | PCI with sirolimus-eluting stents | CABG | 600 | • [48] |  |  |  |  | • [49] |  |
| SYNTAX | Serruys 2009 [50] | International | 2005 – 2007 | MVD or LMCAD | > 50% stenosis of coronary vessel at least 1.5mm in diameter | PCI with TAXUS paclitaxel-eluting stent | CABG | 1800 | • [51] | • [52] | • [53] |  |  | • [54] | • [29] |
| VA CARDS | Kamalesh 2013 [55] | United States | 2006 – 2010 | MVD + Diabetes | Stenosis between 50% and 70% in multivessel disease including left anterior descending coronary artery disease | PCI with DES (any FDA approved) | CABG | 198* | • [55] |  |  |  |  |  |  |

MVD = multi-vessel disease; LMCAD = left main coronary artery disease; FFR = fractional flow reserve; \* terminated early due to slow recruitment; • = present

The sample sizes of the MLTC subgroups ranged between 72 to 953 participants in the PCI and from 69 to 947 in the CABG groups, respectively. Baseline characteristics of included participants, stratified by comorbidities, are presented in Table 2. The proportion of MLTC sample size out of the overall participants in the relevant RCTs were: diabetes (4597/9343, 49.2%), CKD (1068/4580, 24.5%), LVSD (333/3705, 9.0%), female (984/7155, 13.8%), and older age (1197/3001, 39.9%). No studies were identified for PVD or frailty for analysis. In comparison, the baseline characteristics of all trial patients from the 9 included RCTs were: diabetes (4773/10544, 45.3%), CKD (1598/10544, 15.2%), LVSD (8.4%), female (881/10544, 16.7%), older age (1197/10544, 11.4%), and PVD (276/10544, 2.6%); no trials reported participant characteristics by frailty.

**Table 2.** Baseline characteristics of patients, stratified by MLTC.

| Trial | Follow-up (y) | PCI |  |  |  |  |  |  |  |  |  |  |  | CABG |  |  |  |  |  |  |  |  |  |  |  |
| --- | --- | --- | --- | --- | --- | --- | --- | --- | --- | --- | --- | --- | --- | --- | --- | --- | --- | --- | --- | --- | --- | --- | --- | --- | --- |
|  |  | N | Age (y) | Sex (%M) | Hypertension (%) | Dyslipidemia (%) | Previous MI (%) | Stable Angina (%) | Unstable Angina (%) | CKD (%) | Diabetes (%) | LVSD (%) | PVD (%) | N | Age (y) | Sex (%M) | Hypertension (%) | Dyslipidemia (%) | Previous MI (%) | Stable Angina (%) | Unstable Angina (%) | CKD (%) | Diabetes (%) | LVSD (%) | PVD (%) |
| DIABETES |  |  |  |  |  |  |  |  |  |  |  |  |  |  |  |  |  |  |  |  |  |  |  |  |  |
| BEST | 11.8 | 177 | 63.0±8.0 | 118 (66.7) | 121 (68.4) | 99 (55.9) | 5 (2.8) | 79 (44.6) | 77 (43.5) | 6 (3.4) | 177 (100) | 59 (33.3) | 5 (2.8) | 186 | 66±9 | 133 (71.5) | 139 (74.7) | 100 (53.8) | 9 (4.8) | 87 (46.8) | 82 (44.1) | 4 (2.2) | 186 (100) | 60 (32.3) | 9 (4.8) |
| CARDia | 1 | 256 | 64.3 | 181 (70.7) | 196 (76.6) | 237 (92.6) | NR | 201 (78.5) | 55 (21.5) | 14 (5.5) | 256 (100) | 59 (23.0) | 6 (2.3) | 254 | 63.6 | 197 (77.6) | 203 (79.9) | 220 (86.6) | NR | 194 (76.4) | 60 (23.6) | 10 (3.9) | 254 (100) | 79 (31.1) | 13 (5.1) |
| EXCEL | 3 | 286 |  |  |  |  |  |  |  |  |  |  |  | 268 |  |  |  |  |  |  |  |  |  |  |  |
| FAME3 | 3 | 214 | 65.5±7.9 | 169 (79.0) | 185 (86.4) | 174 (81.3) | 66 (30.8) | NR | 72 (33.6) | 23 (10.7) | 214 (100) | 48 (22.4) | NR | 214 | 66.0±7.5 | 174 (81.3) | 199 (93.0) | 183 (85.5) | 71 (33.2) | NR | 76 (35.5) | 22 (10.3) | 214 (100) | 51 (23.8) | NR |
| FREEDOM | 7.5 | 953 | 63.2±8.9 | 698 (73.2) | 806 (84.6) | NR | 250 (26.2) | NR | 304 (31.9) | 73 (7.7) | 953 (100) | 21 (2.2) | NR | 947 | 63.1±9.2 | 658 (69.5) | 806 (85.1) | NR | 237 (25.0) | NR | 279 (29.5) | 56 (5.9) | 947 (100) | 11 (1.2) | NR |
| PRECOMBAT | 10 | 102 | 63.2±8.8 | 72 (70.6) | 67 (65.7) | 51 (50.0) | 3 (2.9) | 45 (44.1) | 51 (50.0) | 2 (2.0) | 102 (100) | 0 | 7 (6.9) | 90 | 63.2±8.7 | 73 (81.1) | 48 (53.3) | 32 (35.6) | 9 (10.0) | 47 (52.2) | 34 (37.8) | 1 (1.1) | 90 (100) | 2 (2.2) | 3 (3.3) |
| SYNTAXES | 10 | 231 | 65.2±9.1 | 165 (71.4) | 172 (74.5) | 185 (80.1) | 76 (32.9) | 129 (55.8) | 74 (32.0) | 42 (18.2) | 231 (100) | 14 (6.1) | 36 (15.6) | 221 | 65.2±9.9 | 156 (70.6) | 144 (65.2) | 177 (80.1) | 67 (30.3) | 121 (54.8) | 60 (27.1) | 43 (19.5) | 221 (100) | 19 (8.6) | 30 (13.6) |
| VA CARDS | 2 | 101 | 62.7±7.1 | 100 (99.0) | 97 (96.0) | NR | 36 (35.6) | 74 (73.3) | 10 (9.9) | 26 (25.7) | 101 (100) | 19 (18.8) | 11 (10.9) | 97 | 62.1±7.4 | 96 (99.0) | 90 (92.8) | NR | 27 (27.8) | 64 (66.0) | 8 (8.2) | 33 (34.0) | 97 (100) | 13 (13.4) | 16 (16.5) |
| CKD |  |  |  |  |  |  |  |  |  |  |  |  |  |  |  |  |  |  |  |  |  |  |  |  |  |
| EXCEL | 3 | 177 | Not extractable |  |  |  |  |  |  |  |  |  |  | 184 | Not extractable |  |  |  |  |  |  |  |  |  |  |
| FREEDOM | 3.8 | 225 | 67.9±7.9 | 141 (62.7) | 213 (94.7) | 189 (84.0) | 63 (28.0) | 147 (65.3) | 57 (25.3) | 225 (100) | NR | NR | 42 (18.7) | 226 | 67.9±8.8 | 144 (63.7) | 210 (92.9) | 191 (84.5) | 53 (23.5) | 159 (70.4) | 58 (25.7) | 226 (100) | NR | NR | 33 (14.6) |
| SYNTAXES | 5 | 158 | 71.9±7.3 | 108 (68.4) | 135 (85.4) | 122 (77.2) | 58 (36.7) | NR | NR | 158 (100) | 44 (27.8) | 3 (1.9) | 21 (13.3) | 151 | 71.6±7.9 | 101 (66.9) | 128 (84.8) | 119 (78.8) | 47 (31.1) | NR | NR | 151 (100) | 50 (33.1) | 8 (5.3) | 24 (15.9) |
| LVSD |  |  |  |  |  |  |  |  |  |  |  |  |  |  |  |  |  |  |  |  |  |  |  |  |  |
| EXCEL | 3 | 43 | 65.0±9.0 | 27 (62.8) | 31 (72.1) | 16 (37.2) | 13 (30.2) | NR | NR | 14 (32.6) | 14 (32.6) | 43 (100) | 11 (25.6) | 31 | 69.8±9.0 | 26 (83.9) | 23 (74.2) | 17 (54.8) | 5 (16.1) | NR | NR | 10 (32.3) | 10 (32.3) | 31 (100) | 3 (9.7) |
| SYNTAXES | 10 | 91 | Not extractable |  |  |  |  |  |  |  |  |  |  | 168 | Not extractable |  |  |  |  |  |  |  |  |  |  |
| SEX (female) |  |  |  |  |  |  |  |  |  |  |  |  |  |  |  |  |  |  |  |  |  |  |  |  |  |
| EXCEL | 3 | 226 | 66.8±10.0 | 226<br>(100) | 183<br>(81.0) | 166<br>(73.5) | NR | 119<br>(52.7) | NR | 62<br>(27.4) | 74<br>(32.7) | 27<br>(11.9) | 24<br>(10.6) | 215 | 67.3±10.4 | 215<br>(100) | 164<br>(76.3) | 161<br>(74.9) | NR | 114<br>(53.0) | NR | 55<br>(25.6) | 62<br>(28.8) | 19<br>(8.8) | 16<br>(7.4) |
| FAME3 | 3 | 141 | 67.6±7.6 | 141<br>(100) | 109<br>(77.3) | 98<br>(69.5) | 50 | NR | 58<br>(41.1) | 4<br>(2.8) | 45<br>(31.9) | 23<br>(16.3) | NR | 124 | 67.7±7.6 | 124<br>(100) | 98<br>(79.0) | 89<br>(71.8) | 45<br>(36.3) | NR | 51<br>(41.1) | 10<br>(8.1) | 40<br>(32.3) | 16<br>(12.9) | NR |
| PRECOMBAT | 10 | 72 | 62.4±9.5 | 72<br>(100) | 41<br>(56.9) | 30<br>(41.7) | 1<br>(1.4) | 35<br>(48.6) | 36<br>(50.0) | 1<br>(1.4) | 30<br>(41.7) | 0 (0) | 5<br>(6.9) | 69 | 63.8±9.6 | 69<br>(100) | 37<br>(53.6) | 28<br>(40.6) | 5<br>(7.2) | 31<br>(44.9) | 35<br>(50.7) | 0<br>(0.0) | 17<br>(24.6) | 0<br>(0.0) | 3<br>(4.3) |
| SYNTAXES | 10 | 213 | 68.3±9.2 | 213<br>(100) | 158<br>(74.2) | 165<br>(77.5) | 65<br>(30.5) | 116<br>(54.5) | 72<br>(33.8) | 62<br>(29.1) | 66<br>(31.0) | 14<br>(6.6) | 25<br>(11.7) | 189 | 67.9±10.0 | 189<br>(100) | 126<br>(66.7) | 149<br>(78.8) | 52<br>(27.5) | 107<br>(56.6) | 63<br>(33.3) | 57<br>(30.2) | 65<br>(34.4) | 8<br>(4.2) | 20<br>(10.6) |
| **OLDER AGE** |  |  |  |  |  |  |  |  |  |  |  |  |  |  |  |  |  |  |  |  |  |  |  |  |  |
| NOBLE (>67) | 3 | 310 | 73 | 243<br>(78.4) | 233<br>(75.2) | 255<br>(82.3) | NR | 255<br>(82.3) | 55<br>(17.7) | NR | 54<br>(17.4) | NR | NR | 312 | 73 | 236<br>(75.6) | 220<br>(70.5) | 249<br>(79.8) | NR | 256<br>(82.1) | 55<br>(17.6) | NR | 54<br>(17.3) | NR | NR |
| SYNTAXES (>70) | 10 | 290 | 75.9±3.6 | 189<br>(65.2) | 209<br>(72.1) | 200<br>(69.0) | 88<br>(30.3) | 156<br>(53.8) | 89<br>(30.7) | 126<br>(43.4) | 67<br>(23.1) | 20<br>(6.9) | 32<br>(11.0) | 285 | 75.6±3.7 | 193<br>(67.7) | 189<br>(66.3) | 198<br>(69.5) | 90<br>(31.6) | 150<br>(52.6) | 91<br>(31.9) | 102<br>(35.8) | 75<br>(26.3) | 17<br>(6.0) | 39<br>(13.7) |

NR = not reported

**Table 3.** Summary of findings, with certainty of finding.

| Certainty assessment |  |  |  |  |  |  |  | No of patients |  | Effect |  | Certainty |
| --- | --- | --- | --- | --- | --- | --- | --- | --- | --- | --- | --- | --- |
| Outcome by MLTC | No of studies | Study design | Risk of bias | Inconsistency | Indirectness | Imprecision | Other cond's | PCI | CABG | Relative (95% CI) | Absolute (95% CI) |  |
| **Mortality (diabetes)** | 8 | randomised trials | not serious<sup>a</sup> | not serious<sup>b</sup> | not serious | not serious | none | 384/2990 (12.8%) | 299/2947 (10.1%) | **RR 1.25**<br>(1.00 to 1.56) | **25 more per 1,000**<br>(from 0 fewer to 57 more) | ⊕⊕⊕⊕<br>High<sup>a,b</sup> |
| **Mortality (CKD)** | 3 | randomised trials | not serious<sup>a</sup> | not serious | not serious | serious<sup>c</sup> | none | 129/560 (23.0%) | 86/561 (15.3%) | **RR 1.48**<br>(1.16 to 1.89) | **74 more per 1,000**<br>(from 25 more to 136 more) | ⊕⊕⊕○<br>Moderate<sup>a,c</sup> |
| **Mortality (LVSD)** | 2 | randomised trials | not serious<sup>a</sup> | not serious | not serious | very serious<sup>c</sup> | none | 47/120 (39.2%) | 40/122 (32.8%) | **RR 1.32**<br>(0.95 to 1.82) | **105 more per 1,000**<br>(from 16 fewer to 269 more) | ⊕⊕○○<br>Low<sup>a,c</sup> |
| **Mortality (female)** | 4 | randomised trials | not serious<sup>a</sup> | not serious | not serious | serious<sup>c</sup> | none | 106/652 (16.3%) | 94/597 (15.7%) | **RR 1.03**<br>(0.81 to 1.31) | **5 more per 1,000**<br>(from 30 fewer to 49 more) | ⊕⊕⊕○<br>Moderate<sup>a,c</sup> |
| **Mortality (older)** | 2 | randomised trials | not serious<sup>a</sup> | not serious | not serious | very serious<sup>c</sup> | none | 164/600 (27.3%) | 156/597 (26.1%) | **RR 1.04**<br>(0.88 to 1.25) | **10 more per 1,000**<br>(from 31 fewer to 65 more) | ⊕⊕○○<br>Low<sup>a,c</sup> |
| **MI (diabetes)** | 6 | randomised trials | not serious<sup>a</sup> | serious<sup>d,e</sup> | not serious | serious<sup>f</sup> | none | 98/1134 (8.6%) | 72/1103 (6.5%) | **RR 1.36**<br>(0.76 to 2.45) | **23 more per 1,000**<br>(from 16 fewer to 95 more) | ⊕⊕○○<br>Low<sup>a,d,f</sup> |
| **MI (CKD)** | 2 | randomised trials | not serious<sup>a</sup> | serious<sup>d,e</sup> | not serious | very serious<sup>c,g</sup> | none | 50/402 (12.4%) | 25/410 (6.1%) | **RR 1.94**<br>(0.65 to 5.85) | **57 more per 1,000**<br>(from 21 fewer to 296 more) | ⊕○○○<br>Very low<sup>a,c,d,g</sup> |
| **MI (LVSD)** | 1 | randomised trials | not serious<sup>a</sup> | serious<sup>e</sup> | not serious | very serious<sup>c,g</sup> | none | 3/43 (7.0%) | 3/31 (9.7%) | **RR 0.72**<br>(0.16 to 3.34) | **27 fewer per 1,000**<br>(from 81 fewer to 226 more) | ⊕○○○<br>Very low<sup>a,c,g</sup> |
| **MI (female)** | 3 | randomised trials | not serious<sup>a</sup> | serious<sup>e</sup> | not serious | very serious<sup>c,g</sup> | none | 36/439 (8.2%) | 25/408 (6.1%) | **RR 1.30**<br>(0.75 to 2.26) | **18 more per 1,000**<br>(from 15 fewer to 77 more) | ⊕○○○<br>Very low<sup>a,c,g</sup> |
| **MI (older)** | 1 | randomised trials | not serious<sup>a</sup> | serious<sup>e</sup> | not serious | very serious<sup>c,g</sup> | none | 33/310 (10.6%) | 12/312 (3.8%) | **RR 2.77**<br>(1.46 to 5.26) | **68 more per 1,000**<br>(from 18 more to 164 more) | ⊕○○○<br>Very low<sup>a,c,g</sup> |
| **Stroke (diabetes)** | 6 | randomised trials | not serious<sup>a</sup> | serious<sup>e</sup> | not serious | serious<sup>f</sup> | none | 25/1134 (2.2%) | 38/1103 (3.4%) | **RR 0.70**<br>(0.42 to 1.17) | **10 fewer per 1,000**<br>(from 20 fewer to 6 more) | ⊕⊕○○<br>Low<sup>a,d,f</sup> |
| **Stroke (CKD)** | 2 | randomised trials | not serious<sup>a</sup> | serious<sup>e</sup> | not serious | very serious<sup>c,f</sup> | none | 14/402 (3.5%) | 26/410 (6.3%) | **RR 0.55**<br>(0.29 to 1.04) | **29 fewer per 1,000**<br>(from 45 fewer to 3 more) | ⊕○○○<br>Very low<sup>a,c,d,f</sup> |
| **Stroke (LVSD)** | 1 | randomised trials | not serious<sup>a</sup> | serious<sup>e</sup> | not serious | very serious<sup>c,g</sup> | none | 2/43 (4.7%) | 1/31 (3.2%) | **RR 1.44**<br>(0.14 to 15.20) | **14 more per 1,000**<br>(from 28 fewer to 458 more) | ⊕○○○<br>Very low<sup>a,c,g</sup> |
| **Stroke (female)** | 3 | randomised trials | not serious<sup>a</sup> | serious<sup>e</sup> | not serious | very serious<sup>c,f</sup> | none | 13/439 (3.0%) | 18/408 (4.4%) | **RR 0.73**<br>(0.36 to 1.49) | **12 fewer per 1,000**<br>(from 28 fewer to 22 more) | ⊕○○○<br>Very low<sup>a,c,d,f</sup> |
| **Stroke (older)** | 1 | randomised trials | not serious<sup>a</sup> | serious<sup>e</sup> | not serious | very serious<sup>c,f</sup> | none | 16/310 (5.2%) | 10/312 (3.2%) | **RR 1.61**<br>(0.74 to 3.49) | **20 more per 1,000**<br>(from 8 fewer to 80 more) | ⊕○○○<br>Very low<sup>a,c,d,f</sup> |
| **Repeat revasc (diabetes)** | 6 | randomised trials | not serious<sup>a</sup> | serious<sup>d</sup> | not serious | not serious<sup>h</sup> | none | 185/1134 (16.3%) | 84/1103 (7.6%) | **RR 2.12**<br>(1.39 to 3.24) | **85 more per 1,000**<br>(from 30 more to 171 more) | ⊕⊕⊕○<br>Moderate<sup>a,d,h</sup> |
| **Repeat revasc (CKD)** | 3 | randomised trials | not serious<sup>a</sup> | not serious | not serious | serious<sup>c,f</sup> | none | 120/560 (21.4%) | 49/561 (8.7%) | **RR 2.45**<br>(1.80 to 3.34) | **127 more per 1,000**<br>(from 70 more to 204 more) | ⊕⊕⊕○<br>Moderate<sup>a,c,f</sup> |
| **Repeat revasc (LVSD)** | 1 | randomised trials | not serious<sup>a</sup> | not serious | not serious | very serious<sup>c,g</sup> | none | 4/43<br>(9.3%) | 2/31<br>(6.5%) | **RR 1.44**<br>(0.28 to 7.38) | **28 more per 1,000**<br>(from 46 fewer to 412 more) | ⊕⊕○○<br>Low<sup>a,c,g</sup> |
| **Repeat revasc (female)** | 2 | randomised trials | not serious<sup>a</sup> | not serious | not serious | very serious<sup>c,g</sup> | none | 46/367<br>(12.5%) | 33/339<br>(9.7%) | **RR 1.28**<br>(0.84 to 1.96) | **27 more per 1,000**<br>(from 16 fewer to 93 more) | ⊕⊕○○<br>Low<sup>a,c,g</sup> |
| **Repeat revasc (older)** | 1 | randomised trials | not serious<sup>a</sup> | not serious | not serious | very serious<sup>c,g</sup> | none | 60/319<br>(18.8%) | 31/312<br>(9.9%) | **RR 1.89**<br>(1.26 to 2.84) | **88 more per 1,000**<br>(from 26 more to 183 more) | ⊕⊕○○<br>Low<sup>a,c,g</sup> |
| **Quality of life (SAQ)** | 2 | randomised trials | not serious<sup>a</sup> | not serious | not serious | very serious<sup>c,g</sup> | publication bias strongly suspected<sup>d</sup> | 1243 | 1232 | - | **MD 0.57 higher**<br>(0.95 lower to 2.09 higher) | ⊕○○○<br>Very low<sup>a,c,g,i</sup> |

CI: confidence interval; RR: risk ratio

### Risk of bias assessment

All the included trials had some concerns of bias due to the lack of blinding, where participants and healthcare professionals delivering the interventions were aware of the assigned intervention. All analyses used intention-to-treat principles. (Figure 2) Additional risk of bias assessments are available in Supplementary Material (Figure S1).

**Figure 2.**
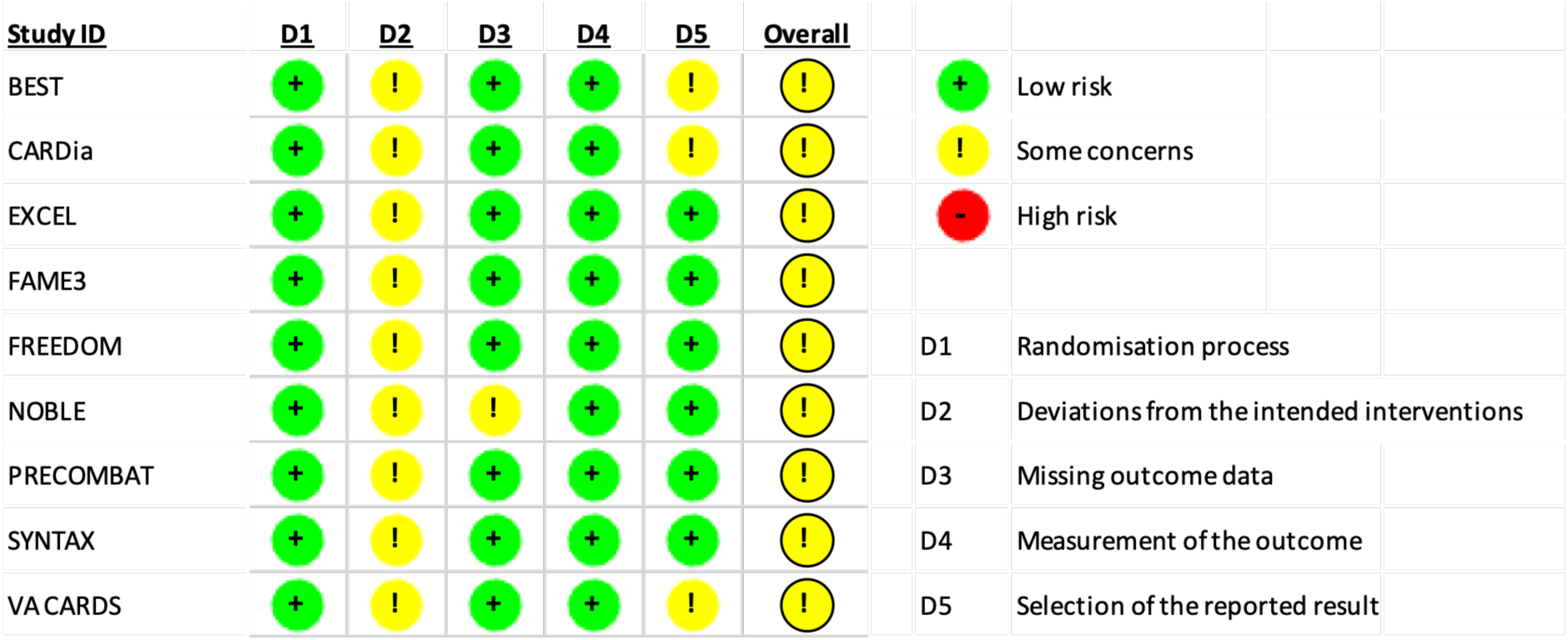
Risk of bias assessment for included RCTs

### Clinical outcomes

Clinical outcomes, including all-cause mortality, MI, stroke, repeat revascularization, and MACCE for MLTC population including diabetes, CKD, LVSD, female, and older age were reported within the 9 included RCTs. All included RCTs used a primary outcome of MACCE, which was a composite outcome using a combination of all-cause mortality, MI, stroke, and repeat revascularisation, although the exact definition varied across RCTs. No studies reported clinical outcomes by PVD or frailty. The median follow-up for diabetes was 5 years (range 1-10 years), CKD 5 years (range 3-5 years), LVSD 7 years (range 3-10 years), female 7 years (range 3-10 years), and older age 7 years (range 3-10 years).

All-cause mortality, stratified by comorbidities, was reported for diabetes (8 studies), CKD (3 studies), LVSD (2 studies), women (4 studies), and older age (2 studies). Mortality was lower following CABG for people with diabetes (RR=1.25, 95%CI: 1.00-1.56), and CKD (RR=1.48, 95%CI: 1.16-1.89). There was no difference in mortality for people with LVSD (RR=1.32, 95%CI: 0.95-1.82). female sex (RR=1.03, 95%CI: 0.81-1.31), or older participants (RR=1.04, 95%CI 0.88-1.25). (Figure 3) When restricted to mortality at 10 years, CABG was favoured for people with diabetes (RR=1.01, 95%CI: 0.83-1.22); no difference was observed for people with LVSD (RR=1.39, 95%CI: 0.99-1.96), female (RR=1.05, 95%CI: 0.79-1.39), or older age (RR=1.06, 95%CI 0.87-1.29), and no study reported 10-year mortality in people with CKD. (Supplementary Material, Figure S2)

**Figure 3.**
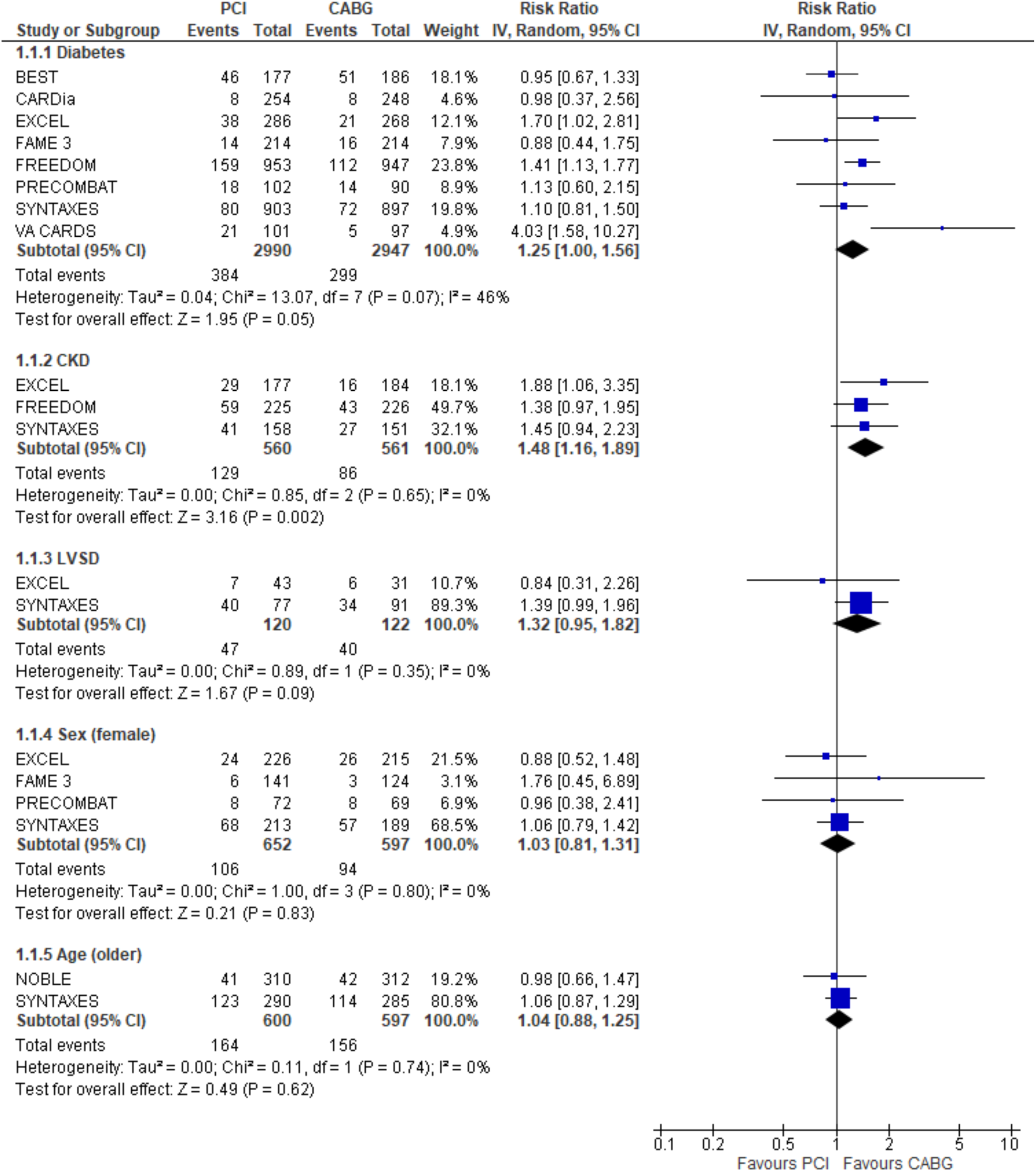
Risk of mortality after CABG versus PCI by MLTC

MI, stratified by comorbidities, was reported for diabetes (6 studies), CKD (2 studies), LVSD (1 study), women (3 studies), and older age (1 study). The risk of MI between PCI or CABG was not different for diabetes (RR=1.36, 95%CI: 0.76-2.45), CKD (RR=1.94, 95%CI 0.65-5.85), LVSD (RR=0.72, 95%CI: 0.16-3.34), or female sex (RR=1.30, 95%CI: 0.75-2.26). The risk of MI was lower with CABG versus PCI for older age (RR=2.77, 95%CI: 1.46-5.26); however, this is based on only 1 RCT. (Figure 4)

**Figure 4.**
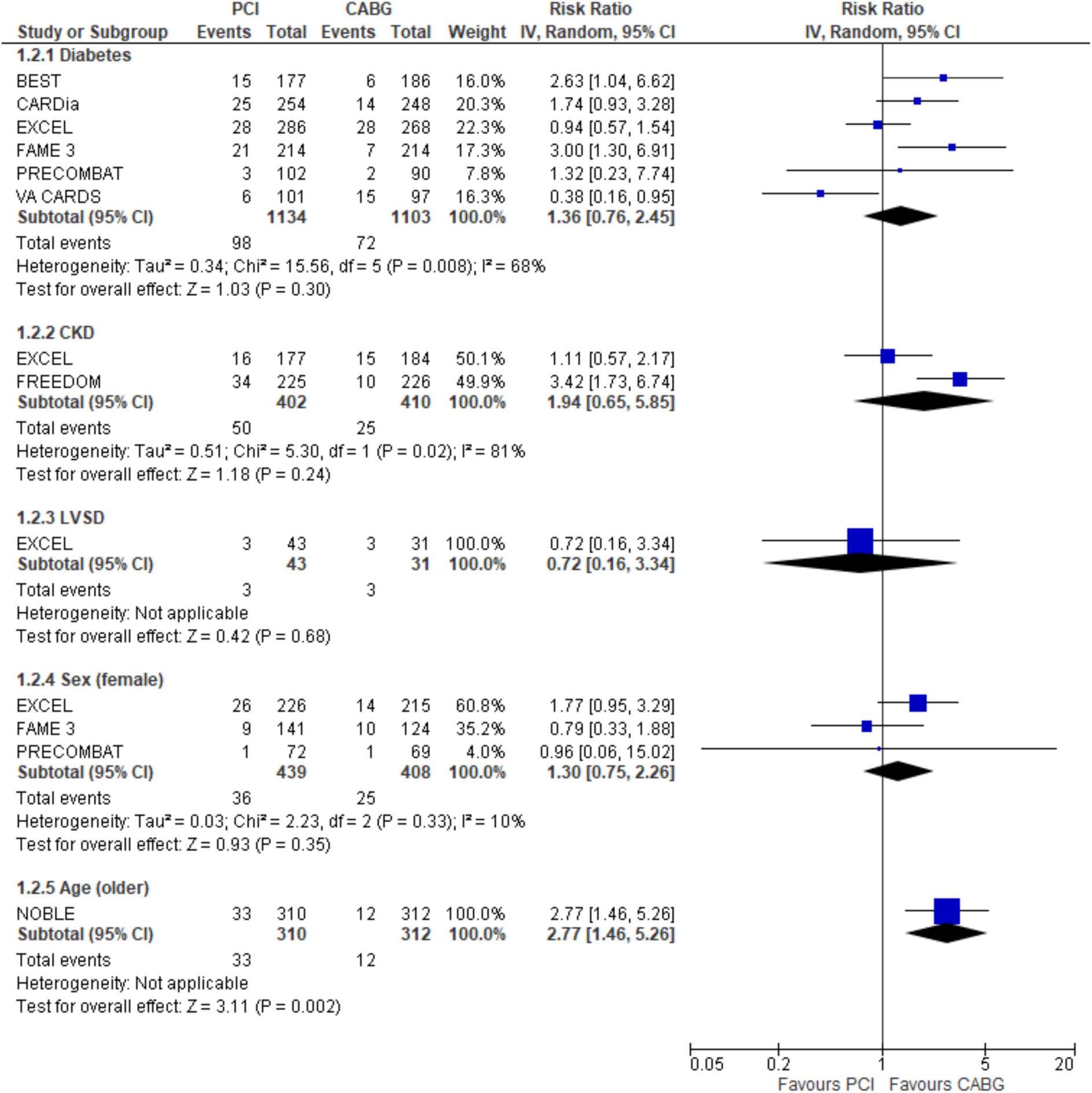
Risk of MI after CABG versus PCI by MLTC

Stroke, stratified by comorbidities, was reported for diabetes (6 studies), CKD (2 studies), LVSD (1 study), women (3 studies), and older age (1 study). There was no difference in stroke events for diabetes (RR=0.70, 95%CI: 0.42-1.17), CKD (RR=0.55, 95%CI: 0.29-1.04), LVSD (RR=1.44, 95%CI: 0.14-15.20), female sex (RR=0.73, 95%CI: 0.36-1.49), or older age (RR=1.61, 95%CI:0.74-3.49). (Figure 5)

**Figure 5.**
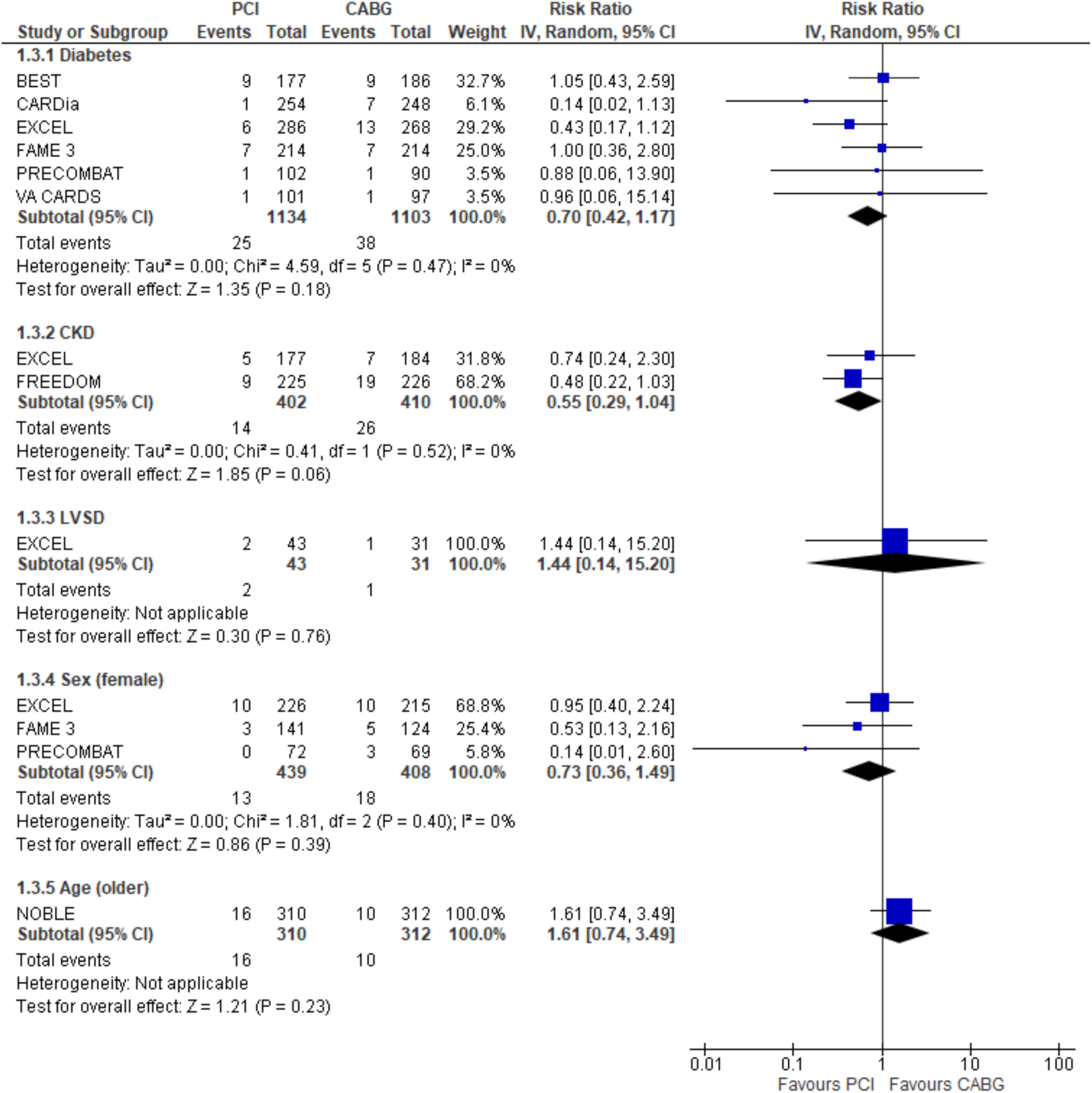
Risk of stroke after CABG versus PCI by MLTC

Repeat revascularisation, stratified by comorbidities, was reported for diabetes (6 studies), CKD (3 studies), LVSD (1 study), women (2 studies), and older age (1 study). CABG reduced repeat revascularisation for people with diabetes (RR=2.12, 95%CI: 1.39-3.24), CKD (RR=2.45, 95%CI 1.80-3.34), and older age (RR=1.89, 95%CI: 1.26-2.84). There was no difference in repeat revascularisation for people with LVSD (RR=1.44, 95%CI: 0.28-7.38), or female sex (RR=1.28, 95%CI: 0.84-1.96). (Figure 6)

**Figure 6.**
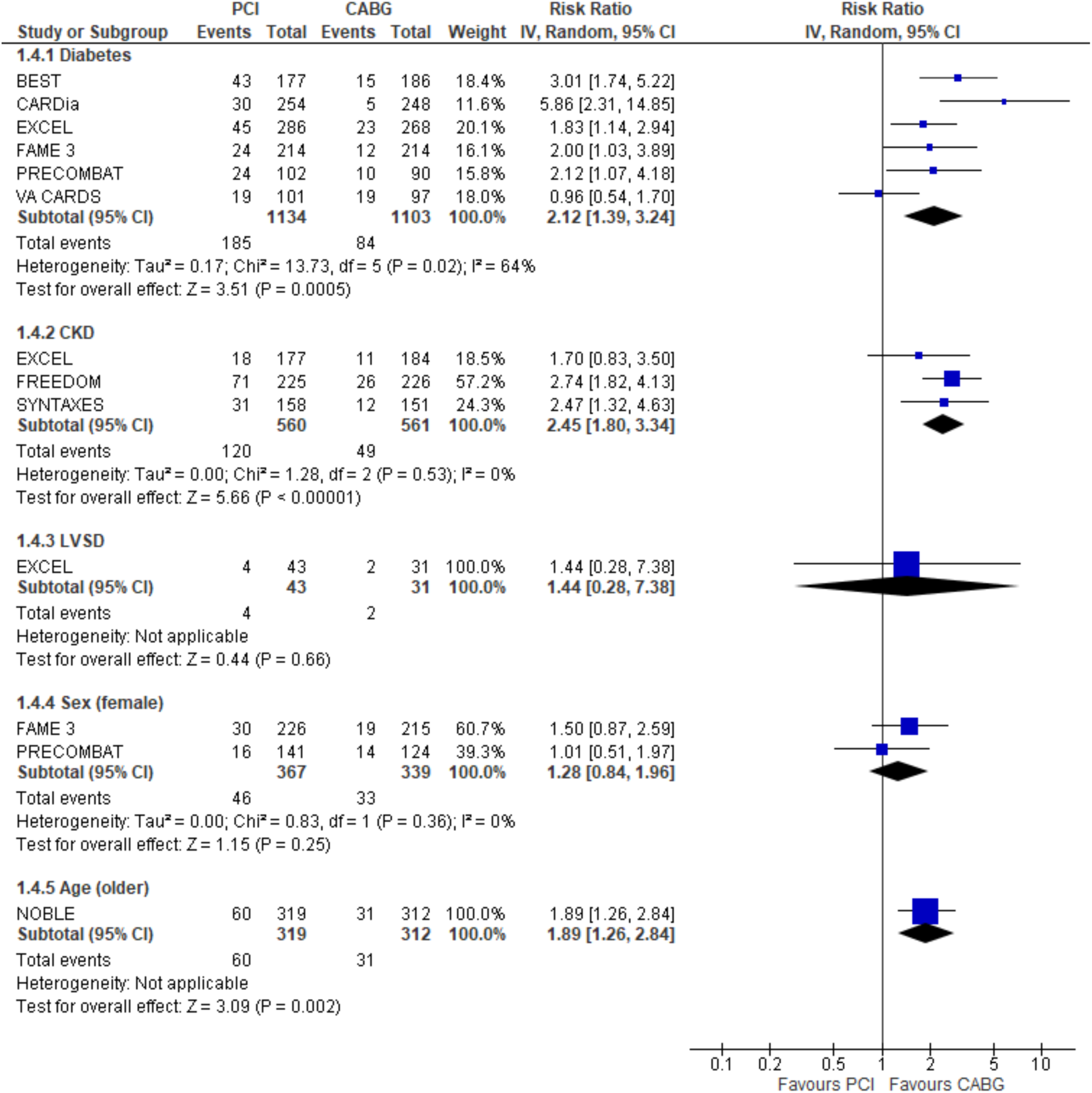
Risk of repeat revascularisation after CABG versus PCI by MLTC

MACCE, stratified by comorbidities, was reported for diabetes (6 studies), CKD (3 studies), LVSD (1 study), women (3 studies), and older age (2 studies). CABG reduced the risk of MACCE in people with CKD (RR=1.38, CI% 1.15-1.67) and older age (RR=1.38, 95%CI: 1.05-1.81). There was no difference in MACCE events for diabetes (RR=1.18, 95%CI: 0.99-1.49), LVSD (RR=0.99, 95%CI: 0.45-2.17), or female (RR=1.16, 95%CI: 0.88-1.54). (Figure 7)

**Figure 7.**
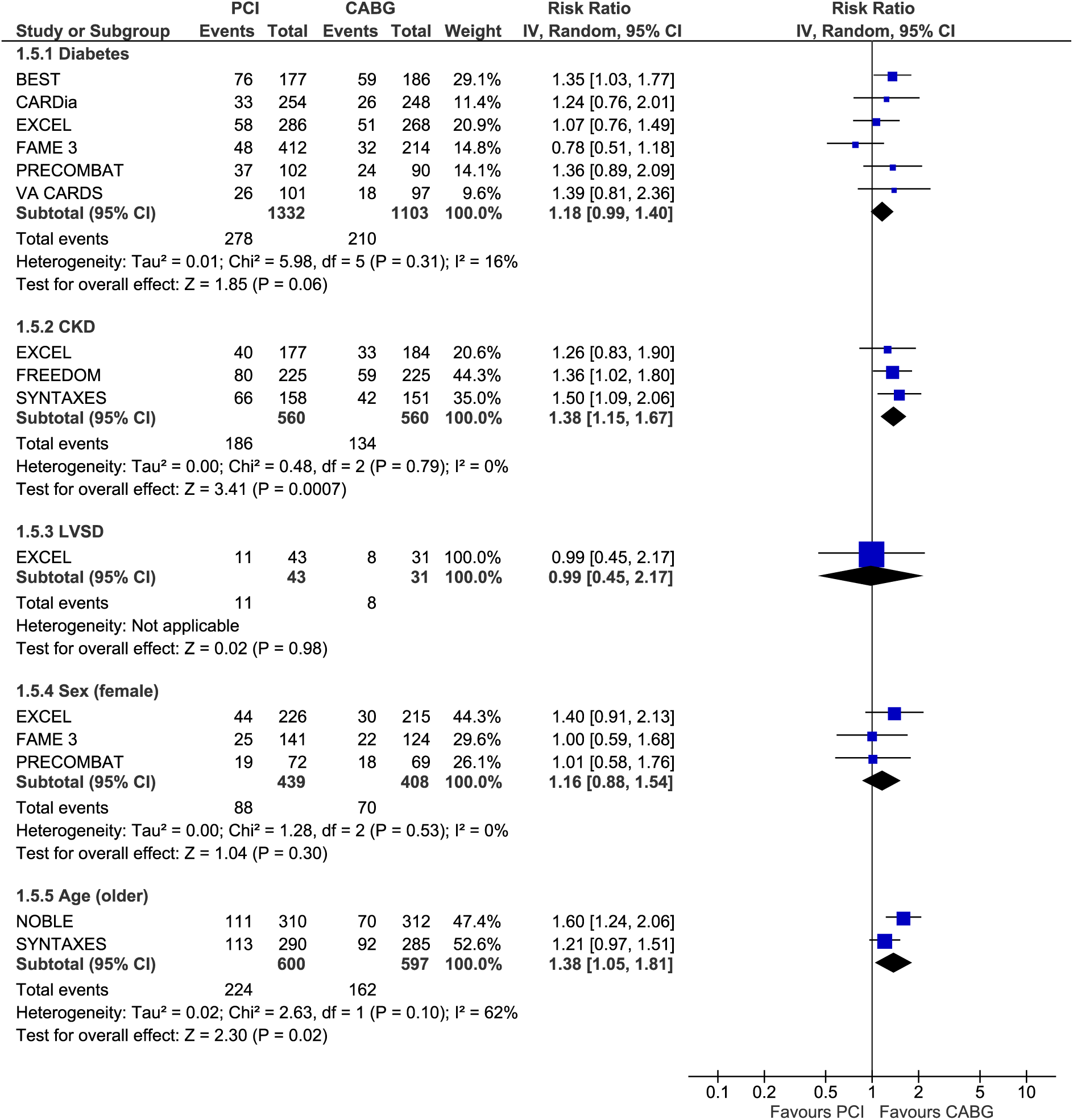
Risk of MACCE after CABG versus PCI by MLTC

### Quality of life

Only two studies reported quality of life outcomes stratified by comorbidities: FREEDOM, as the entire trial population included diabetes; and SYNTAX, which stratified by older age. The common quality of life measure used was the Seattle Angina Questionnaire (SAQ). The results showed that CABG reduced angina frequency (MD=-1.51, 95%CI: -2.97--0.04), but there was no difference in physical limitation (MD=2.42, 95%CI: -1.59-6.43), treatment satisfaction (MD=1.30, 95%CI:-1.40-4.04), or quality of life (MD=1.39, 95%CI:=-1.88-4.67) at 5 years. (Figure 8)

**Figure 8.**
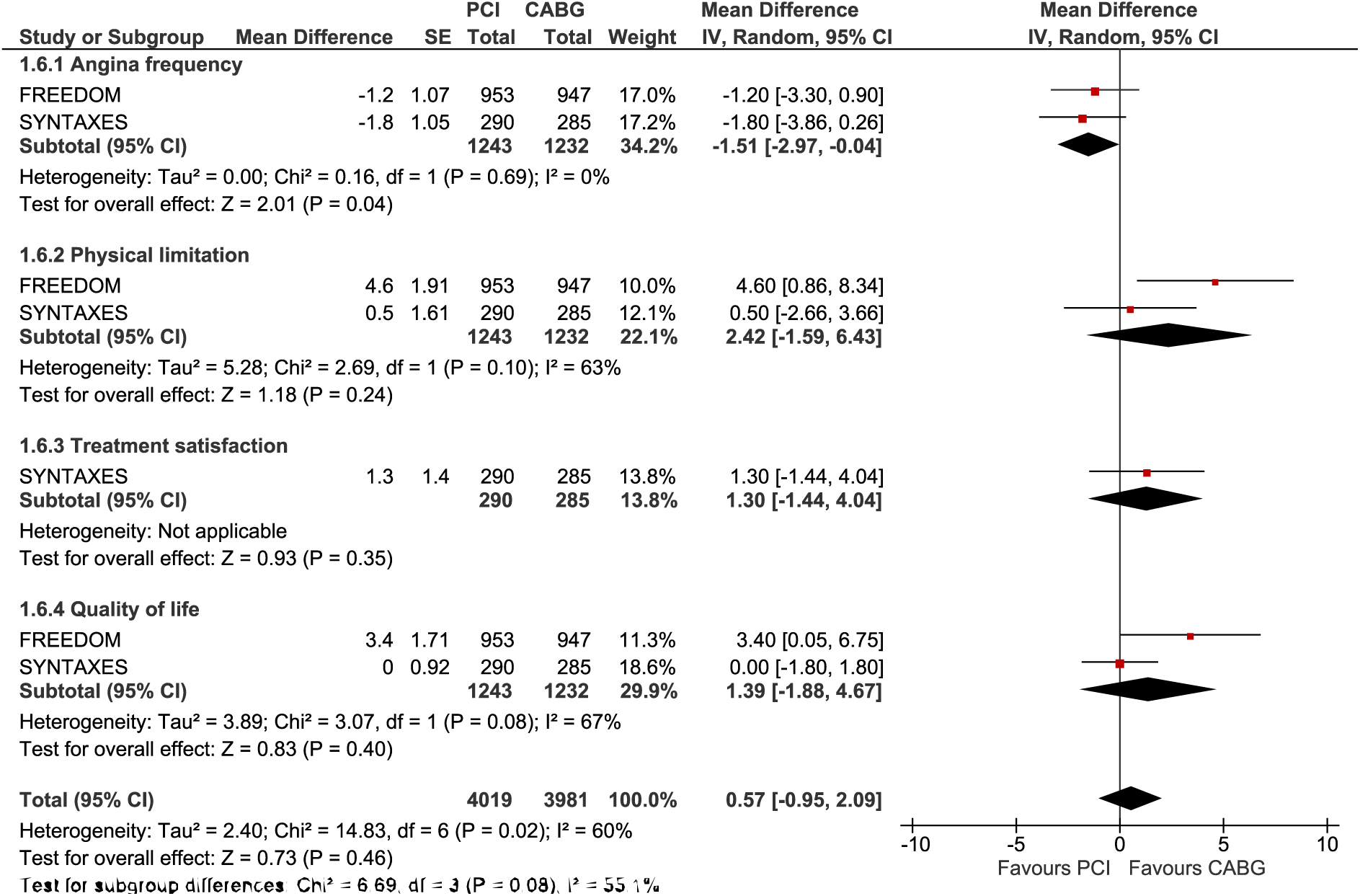
Quality of life after CABG versus PCI by MLTC

### Grading of Recommendations Assessment, Development and Evaluation

The certainty of findings, assessed through the approach, is predominantly low and very low for all outcomes, with the exception of diabetes where certainty was High for all-cause mortality and Moderate for repeat revascularisation. (Table 4)

## DISCUSSION

This systematic review and meta-analysis of 9 trials demonstrated a significant knowledge gap with respect to the effects of CABG versus PCI in people with MLTC, specifically diabetes, CKD, LVSD, PVD, frailty, female, and older age. The certainty of the evidence was generally low to very low, due to the imprecision of the estimates and the number of the included studies; the exception was diabetes, which is reflected in major clinical guidelines as Class I Level A/B evidence, where a revascularisation strategy with CABG is preferred in people with MVD.[3, 4]

### Diabetes

People with diabetes undergoing CABG versus PCI have reduced risk of mortality and repeat revascularisation, with no difference in MI, stroke, or MACCE. This is consistent with previous meta-analyses. For example, Head et al. included 11 RCTs in a pooled analysis of individual patient data (IPD) and estimated a greater 5-year all-cause mortality in diabetic patients with MVD (15.5% after PCI vs. 10.0% after CABG; HR 1.48, 95% CI 1.19-1.84). They reported no difference in mortality for patients with LMCAD regardless of diabetes status.[17] This is also supported by a separate IPD analysis of 4 RCTs specifically examining LMCAD and diabetes, which found no difference in the risk of death, a higher risk of stroke, lower risk of spontaneous MI and lower risk of repeat revascularisation with CABG.[18] Separate analyses for MVD and LMCAD was not possible in this review.

### CKD

CABG reduces the risk of mortality, repeat revascularisation, and MACCE, with no difference in the risk of MI or stroke. This contrasts with the IPD analysis by Charytan et al. in which they showed that CABG reduces the risk of MI and revascularisation, with no difference in mortality between PCI or CABG in those with CKD.[19] However, only 3 of the 10 RCTs included in that analyses enrolled after 2000, and some of the RCTs, such as BARI and MASS, compared invasive strategies, either PCI or CABG, against a conservative strategy.

### LVSD

There was limited evidence with regards to revascularisation choices in people with LVSD, as our study only found 1 to 2 eligible RCTs depending on the clinical outcome. This resulted in low or very low certainty assessments due in large part by wide confidence intervals. An existing reconstructed IPD analysis consisting of 3 RCTs and 11 observational studies by Lee et al. suggested that PCI was associated with higher risk of all-cause mortality, repeat revascularisation and MI, but lower risk of short-term stroke.[20] Unfortunately, RCTs represented only 1.3% of the total patient numbers included, and it is likely indication bias has skewed the results from true estimates. These results support the rationale for the ongoing STICH3-BCIS4 trial.[21]

### PVD or frailty

Our study did not identify any RCTs or subgroup analyses specifically considering PVD or frailty. During title and abstract screening, an abstract reporting a substudy on people with PVD in the EXCEL trial was identified, which suggested no difference in all-cause death, MI, and stroke, but higher rate of ischemia-driven revascularisation at 3 years; more granular comparison between PCI and CABG was not available.[22] No studies reported the effect of frailty on the treatment efficacy between PCI and CABG.

### Female

There was no difference in the risk of mortality, MI, stroke, repeat revascularisation, or MACCE for CABG versus PCI for female sex. Interestingly, Sotomi et al. suggested that CABG is more favourable in Western women based on an IPD analysis of 3 trials, noting the presence of heterogeneous sex-treatment interaction.[23] Similar study level meta-analysis including 6 RCTs also found similar results for MVD, although no full-text publication is available and only a composite outcome of all-cause death, MI, and/or stroke was reported;[24] no differences were found for LMCAD.[25] Sex-based differences in revascularisation outcome remains an important question, and the ongoing RECHARGE:W trial may address this uncertainty.[26]

### Older age

There was limited evidence for people with older age, and searches only identified 1 to 2 eligible RCTs depending on the clinical outcome. Furthermore, there was substantial heterogeneity in the age cut-off across RCTs. In an IPD analysis of 3 RCTs, Chang et al. reported that CABG did not show a mortality benefit over PCI, although there was a benefit for the composite outcome of death or myocardial infarction in adults aged 70 to 89 years.[27] This contrasts with Head et al. subgroup analysis showing reduced risk of mortality after CABG at 5 years for those aged ≥ 65 years old.[17] As the number of older adults grows rapidly worldwide, it is imperative that future studies consider the aging population whilst acknowledging that age is an arbitrary construct.

### Quality of life

There is limited data on the quality of life after revascularisation for MLTC populations. Quality of life outcomes were reported by the complete trial population and not by subgroups. In our study, quality of life data was available only for diabetes from FREEDOM[28] and age from SYNTAX[29], therefore limiting the generalisability of the result. In a wider systematic review and meta-analysis, Dimagli et al. reported both PCI and CABG led to marked improvements in quality of life, and that any differences between the two procedures were modest in comparison.[30]

Whilst the above discussion highlights individual MLTC, it is important to note that these conditions do not exist in isolation. Baseline characteristics of all the included studies report greater comorbidities in those with MLTC than those without. The interaction in the real-world population, for example CKD patients who may also have diabetes, frailty and are older, is therefore not truly reflected in the RCT population, limiting the generalisability of results to the target population.

### Limitations

There are several limitations to this review. First, there is a lack of high-quality evidence reflected by the number of included studies and the GRADE certainty assessment. With the exception of diabetes, this review found very little evidence to support revascularisation decision in people with MLTC.

Second, PCI has evolved in the last two decades with secondary and tertiary generation stents, routine physiological assessment of coronary lesions and intravascular imaging, making true comparison between the long-term effectiveness of PCI and CABG challenging.

Third, there is notable heterogeneity between RCTs. For instance, MVD and LMCAD are considered distinct subsets of CAD yet there is insufficient evidence for separate analysis in CABG vs PCI, let alone MLTC considerations. Different anatomical complexity, SYNTAX score, patient selection, definition of eligibility and clinical outcomes, and follow-up duration are all factors which cannot be addressed in this review.

Fourth, almost all the results included in this review are reported subgroup analyses, whether pre-defined or post-hoc, further highlighting the evidence gap. With the exception of diabetes, our results should be considered exploratory and hypothesis generating.

Finally, this review only considers the choice of PCI and CABG for revascularisation and makes no comparison between conservative versus invasive treatment, or optimal medical therapy vs. CABG.

## Conclusion

There is a substantial knowledge gap in the existing evidence with respect to the effects of CABG versus PCI on clinical and patient reported outcomes in high-risk groups. More robust clinical trials which evaluate MLTC populations are necessary for better evidence-informed and personalised care.

## TRANSPARENCY STATEMENT

The lead author affirms that this manuscript is an honest, accurate, and transparent account of the study being reported; that no important aspects of the study have been omitted; and that any discrepancies from the study as planned (and, if relevant, registered) have been explained.

## Data Availability

All data produced in the present study are available upon reasonable request to the authors

