## Supplementary material for "Coronary revascularisation in people with multiple long-term conditions: a systematic review and meta-analysis"

**Table S1**. Search strategy on Ovid MEDLINE(R)
*This is the search strategy for the effectiveness part of the mixed methods systematic review; a separate search strategy was used for the experience aspect (not included here).*

1 exp Percutaneous Coronary Intervention/ or Stents/ or Angioplasty/ or ("Percutaneous revasculari*ation" or "percutaneous transluminal coronary angioplast*" or "coronary angioplast*" or PCI or PTCA or PPCI or (percutaneous coronary adj2 (intervention* or revasculari*ation*)) or (balloon adj2 (coronary or dilat* or angioplast*)) or (coronary adj2 stent*)).ti,ab. 183552
2 exp Coronary Artery Bypass/ or ("Coronary Artery Bypass" or "CABG" or "coronary artery bypass graft*" or "aortocoronary bypass" or "bypass surgery" or "surgical revasculari*ation").ti,ab. 90721
3 exp conservative management/ or (("Conservative" or "medical" or "optim*") adj2 ("strateg*" or "treatment*" or "therapy")).ti,ab. 253214
4 (1 and 2) or (1 and 3) or (2 and 3) 26563
5 exp Myocardial revascularization/ or ("coronary reperfusion" or (coronary adj2 revasculari*ation)).ti,ab. 105761
6 exp Coronary artery disease/ or Myocardial ischemia/ 121008
7 ("coronary artery disease" or "myocardial isch*emia" or "cardiovascular disease").ti,ab. 304794
8 5 or 6 or 7 448059
9 exp Comorbidity/ or Multimorbidity/ or Chronic Disease/ 415536
10 ("multiple chronic conditions" or "multiple co?morbid*" or "multiple long term conditions" or "multiple long-term conditions").ti,ab. 7955
11 (multimorbid* or multi-morbid* or comorbid* or co-morbid* or polymorbid* or poly-morbid* or polychronic* or multidisease* or multi-disease*).ti,ab. 303545
12 ((multip* or coexist* or co-exist* or coocur* or co-occur* or concurrent or chronic or persistent or long*term) adj2 (illness* or disease* or condition* or disorder* or syndrom*)).ti,ab. 454010
13 exp Aged/ or Frail Elderly/ or frailty/ or Peripheral arterial disease/ or diabetes mellitus/ or kidney failure, chronic/ or kidney diseases/ or renal insufficiency/ or heart failure/ or stroke volume/ 4031980
14 (Elder* or frail* or "peripheral (vascular OR arter*) adj2 (disease* OR condition* OR disorder*) OR diabetes OR chronic kidney disease OR heart failure OR ejection fraction*").ti,ab. 353456
15 9 or 10 or 11 or 12 or 13 or 14 4830596
16 4 and 8 and 15 9921
17 (exp randomized controlled trial/ or controlled clinical trial.pt. or randomized.ab. or placebo.ab. or drug therapy.fs. or randomly.ab. or trial.ab. or groups.ab. NOT (exp animals/ not humans.sh.) 5410733
18 Epidemiologic studies/ or exp case control studies/ or exp cohort studies/ or Case control.tw. or (cohort adj (study or studies)).tw. or Cohort analy$.tw. or (follow up adj (study or studies)).tw. or (observational adj (study or studies)).tw. or Longitudinal.tw. or retrospective.tw. or cross sectional.tw. or cross-sectional studies/ 4174750
19 16 and (17 or 18) 6890

**Table S2. PRISMA checklist**

| Section and Topic | Item # | Checklist item | Location where item is reported |
|---|---|---|---|
| **TITLE** | | | |
| Title | 1 | Identify the report as a systematic review. | Page 1 |
| **ABSTRACT** | | | |
| Abstract | 2 | See the PRISMA 2020 for Abstracts checklist. | Page 2 |
| **INTRODUCTION** | | | |
| Rationale | 3 | Describe the rationale for the review in the context of existing knowledge. | Page 3 |
| Objectives | 4 | Provide an explicit statement of the objective(s) or question(s) the review addresses. | Page 3 |
| **METHODS** | | | |
| Eligibility criteria | 5 | Specify the inclusion and exclusion criteria for the review and how studies were grouped for the syntheses. | Page 4 |
| Information sources | 6 | Specify all databases, registers, websites, organisations, reference lists and other sources searched or consulted to identify studies. Specify the date when each source was last searched or consulted. | Page 4 |
| Search strategy | 7 | Present the full search strategies for all databases, registers and websites, including any filters and limits used. | Supp page 1 |
| Selection process | 8 | Specify the methods used to decide whether a study met the inclusion criteria of the review, including how many reviewers screened each record and each report retrieved, whether they worked independently, and if applicable, details of automation tools used in the process. | Page 5 |
| Data collection process | 9 | Specify the methods used to collect data from reports, including how many reviewers collected data from each report, whether they worked independently, any processes for obtaining or confirming data from study investigators, and if applicable, details of automation tools used in the process. | Page 5 |
| Data items | 10a | List and define all outcomes for which data were sought. Specify whether all results that were compatible with each outcome domain in each study were sought (e.g. for all measures, time points, analyses), and if not, the methods used to decide which results to collect. | Page 5 |
| | 10b | List and define all other variables for which data were sought (e.g. participant and intervention characteristics, funding sources). Describe any assumptions made about any missing or unclear information. | Page 5 |
| Study risk of bias assessment | 11 | Specify the methods used to assess risk of bias in the included studies, including details of the tool(s) used, how many reviewers assessed each study and whether they worked independently, and if applicable, details of automation tools used in the process. | Page 5 |
| Effect measures | 12 | Specify for each outcome the effect measure(s) (e.g. risk ratio, mean difference) used in the synthesis or presentation of results. | Page 5 |
| Synthesis methods | 13a | Describe the processes used to decide which studies were eligible for each synthesis (e.g. tabulating the study intervention characteristics and comparing against the planned groups for each synthesis (item #5)). | Page 5 |
| | 13b | Describe any methods required to prepare the data for presentation or synthesis, such as handling of missing summary statistics, or data conversions. | N/A |
| | 13c | Describe any methods used to tabulate or visually display results of individual studies and syntheses. | Page 5 |
| | 13d | Describe any methods used to synthesize results and provide a rationale for the choice(s). If meta-analysis was performed, describe the model(s), method(s) to identify the presence and extent of statistical heterogeneity, and software package(s) used. | Page 5 |
| | 13e | Describe any methods used to explore possible causes of heterogeneity among study results (e.g. subgroup analysis, meta-regression). | Page 5 |
| | 13f | Describe any sensitivity analyses conducted to assess robustness of the synthesized results. | N/A |
| Reporting bias | 14 | Describe any methods used to assess risk of bias due to missing results in a synthesis (arising from reporting biases). | Page 5 |

| Section and Topic | Item # | Checklist item | Location where item is reported |
|---|---|---|---|
| assessment | | | |
| Certainty assessment | 15 | Describe any methods used to assess certainty (or confidence) in the body of evidence for an outcome. | Page 5 |
| **RESULTS** | | | |
| Study selection | 16a | Describe the results of the search and selection process, from the number of records identified in the search to the number of studies included in the review, ideally using a flow diagram. | Page 6 |
| | 16b | Cite studies that might appear to meet the inclusion criteria, but which were excluded, and explain why they were excluded. | Page 6 |
| Study characteristics | 17 | Cite each included study and present its characteristics. | Page 14 |
| Risk of bias in studies | 18 | Present assessments of risk of bias for each included study. | Page 20 |
| Results of individual studies | 19 | For all outcomes, present, for each study: (a) summary statistics for each group (where appropriate) and (b) an effect estimate and its precision (e.g. confidence/credible interval), ideally using structured tables or plots. | Page 7 |
| Results of syntheses | 20a | For each synthesis, briefly summarise the characteristics and risk of bias among contributing studies. | Page 7 |
| | 20b | Present results of all statistical syntheses conducted. If meta-analysis was done, present for each the summary estimate and its precision (e.g. confidence/credible interval) and measures of statistical heterogeneity. If comparing groups, describe the direction of the effect. | Page 7 |
| | 20c | Present results of all investigations of possible causes of heterogeneity among study results. | Page 7 |
| | 20d | Present results of all sensitivity analyses conducted to assess the robustness of the synthesized results. | N/A |
| Reporting biases | 21 | Present assessments of risk of bias due to missing results (arising from reporting biases) for each synthesis assessed. | Page 7 |
| Certainty of evidence | 22 | Present assessments of certainty (or confidence) in the body of evidence for each outcome assessed. | Page 9 |
| **DISCUSSION** | | | |
| Discussion | 23a | Provide a general interpretation of the results in the context of other evidence. | Page 22 |
| | 23b | Discuss any limitations of the evidence included in the review. | Page 24 |
| | 23c | Discuss any limitations of the review processes used. | Page 24 |
| | 23d | Discuss implications of the results for practice, policy, and future research. | Page 22 |
| **OTHER INFORMATION** | | | |
| Registration and protocol | 24a | Provide registration information for the review, including register name and registration number, or state that the review was not registered. | Page 9 |
| | 24b | Indicate where the review protocol can be accessed, or state that a protocol was not prepared. | Page 4 |
| | 24c | Describe and explain any amendments to information provided at registration or in the protocol. | N/A |
| Support | 25 | Describe sources of financial or non-financial support for the review, and the role of the funders or sponsors in the review. | Page 27 |
| Competing interests | 26 | Declare any competing interests of review authors. | Page 27 |
| Availability of data, code and | 27 | Report which of the following are publicly available and where they can be found: template data collection forms; data extracted from included studies; data used for all analyses; analytic code; any other materials used in the review. | N/A |

| Section and Topic | Item # | Checklist item | Location where item is reported |
|---|---|---|---|
| other materials | | | |

**Figure S1**. Risk of bias assessment for included subgroup studies

| Study ID | First Author & Year | D1 | D2 | D3 | D4 | D5 | Overall |
|---|---|---|---|---|---|---|---|
| BEST_diabetes | Kim 2023 | + | ! | ! | + | + | ! |
| EXCEL_diabetes | Milojevic 2019 | + | ! | + | + | + | ! |
| FAME3_diabetes | Takahashi 2025 | + | ! | + | + | + | ! |
| PRECOMBAT_diabetes | Jeong 2021 | + | ! | + | + | + | ! |
| SYNTAXES_diabetes | Wang 2021 | + | ! | ! | + | + | ! |
| EXCEL_CKD | Giustino 2018 | + | ! | + | + | + | ! |
| FREEDOM_CKD | Baber 2016 | + | ! | + | + | + | ! |
| SYNTAXES_CKD | Milojevic 2018 | + | ! | + | + | ! | ! |
| EXCEL_LVSD | Thujis 2020 | + | ! | + | + | + | ! |
| SYNTAXES_LVSD | Masuda 2024 | + | ! | + | + | ! | ! |
| EXCEL_female | Serruys 2018 | + | ! | + | + | ! | ! |
| FAME3_female | Takahashi 2025 | + | ! | + | + | + | ! |
| PRECOMBAT_female | Yang 2022 | + | ! | + | + | + | ! |
| SYNTAXES_female | Hara 2020 | + | ! | + | + | + | ! |
| NOBLE_age | Steigen 2021 | + | ! | + | + | ! | ! |
| SYNTAX_age | Ono 2020 | + | ! | + | + | + | ! |
| FREEDOM_QoL | Abdallah 2013 | + | ! | + | + | + | ! |

| | |
|---|---|
| + | Low risk |
| ! | Some concerns |
| - | High risk |
| D1 | Randomisation process |
| D2 | Deviations from the intended ir |
| D3 | Missing outcome data |
| D4 | Measurement of the outcome |
| D5 | Selection of the reported result |

**Figure S2**. Risk of 10-year mortality after CABG versus PCI by MLTC

| Study or Subgroup | PCI Events | PCI Total | CABG Events | CABG Total | Weight | Risk Ratio IV, Random, 95% CI |
|---|---|---|---|---|---|---|
| **1.7.1 Diabetes** | | | | | | |
| BEST | 46 | 177 | 51 | 186 | 33.0% | 0.95 [0.67, 1.33] |
| PRECOMBAT | 18 | 102 | 14 | 90 | 9.4% | 1.13 [0.60, 2.15] |
| SYNTAXES | 80 | 231 | 72 | 212 | 57.6% | 1.02 [0.79, 1.32] |
| **Subtotal (95% CI)** | | **510** | | **488** | **100.0%** | **1.01 [0.83, 1.22]** |
| Total events | 144 | | 137 | | | |
| Heterogeneity: Tau² = 0.00; Chi² = 0.26, df = 2 (P = 0.88); I² = 0% | | | | | | |
| Test for overall effect: Z = 0.05 (P = 0.96) | | | | | | |
| **1.7.2 CKD** | | | | | | |
| **Subtotal (95% CI)** | | **0** | | **0** | | **Not estimable** |
| Total events | 0 | | 0 | | | |
| Heterogeneity: Not applicable | | | | | | |
| Test for overall effect: Not applicable | | | | | | |
| **1.7.3 LVSD** | | | | | | |
| SYNTAXES | 40 | 77 | 34 | 91 | 100.0% | 1.39 [0.99, 1.96] |
| **Subtotal (95% CI)** | | **77** | | **91** | **100.0%** | **1.39 [0.99, 1.96]** |
| Total events | 40 | | 34 | | | |
| Heterogeneity: Not applicable | | | | | | |
| Test for overall effect: Z = 1.89 (P = 0.06) | | | | | | |
| **1.7.4 Sex (female)** | | | | | | |
| PRECOMBAT | 8 | 72 | 8 | 69 | 9.1% | 0.96 [0.38, 2.41] |
| SYNTAXES | 68 | 213 | 57 | 189 | 90.9% | 1.06 [0.79, 1.42] |
| **Subtotal (95% CI)** | | **285** | | **258** | **100.0%** | **1.05 [0.79, 1.39]** |
| Total events | 76 | | 65 | | | |
| Heterogeneity: Tau² = 0.00; Chi² = 0.04, df = 1 (P = 0.84); I² = 0% | | | | | | |
| Test for overall effect: Z = 0.34 (P = 0.74) | | | | | | |
| **1.7.5 Age (older)** | | | | | | |
| SYNTAXES | 123 | 290 | 114 | 285 | 100.0% | 1.06 [0.87, 1.29] |
| **Subtotal (95% CI)** | | **290** | | **285** | **100.0%** | **1.06 [0.87, 1.29]** |
| Total events | 123 | | 114 | | | |
| Heterogeneity: Not applicable | | | | | | |
| Test for overall effect: Z = 0.59 (P = 0.56) | | | | | | |

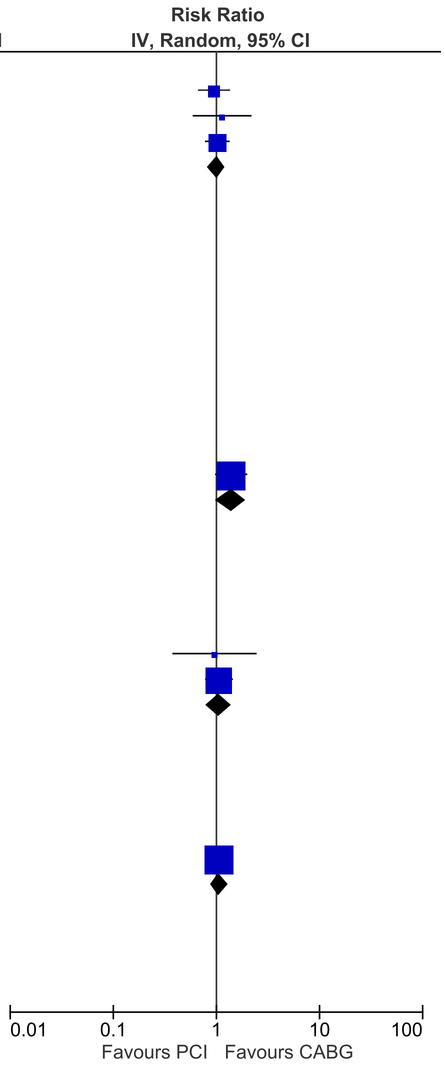
